# Demographic and Clinicopathologic Risk Factors for Colorectal Adenoma Recurrence: A Large-Scale Surveillance Cohort Study of 59,667 Adults

**DOI:** 10.1101/2025.03.28.25324826

**Authors:** Usman Ayub Awan, Qingyuan Song, Kristen K. Ciombor, Adetunji T. Toriol, Jungyoon Choi, Timothy Su, Xiao-ou Shu, Kamran Idrees, Kay M. Washington, Wei Zheng, Wanqing Wen, Zhijun Yin, Xingyi Guo

**Affiliations:** Division of Epidemiology, Department of Medicine, Vanderbilt-Ingram Cancer Center, Vanderbilt University Medical Center, Nashville, TN, USA; Department of Medical Laboratory Technology, The University of Haripur, Haripur, Khyber Pakhtunkhwa, Pakistan; Department of Biomedical Informatics, Vanderbilt University Medical Center, Nashville, TN, USA; Department of Department of Computer Science, Vanderbilt University, Nashville, TN, USA; Division of Hematology/Oncology, Department of Medicine, Vanderbilt University Medical Center, Vanderbilt-Ingram Cancer Center, Nashville, TN, USA; Division of Public Health Sciences, Department of Surgery, and Siteman Cancer Center, Washington University School of Medicine St. Louis, MO, USA; Division of Oncology and Hematology, Department of Internal Medicine, Korea University Ansan Hospital, Korea University College of Medicine, Ansan, Korea; Division of Surgical Oncology and Endocrine Surgery, Department of Surgery, Vanderbilt University Medical Center, Vanderbilt-Ingram Cancer Center Nashville, TN, USA; Department of Pathology, Vanderbilt University Medical Center, Nashville, Tennessee; Department of Electrical and Computer Engineering, Vanderbilt University, Nashville, TN, USA

**Keywords:** Colorectal Adenoma Recurrence, Demographic Disparities, Surveillance, Poly Characteristics, Cohort Study

## Abstract

**Background:** Current colorectal surveillance guidelines emphasize adenoma characteristics but overlook temporal, racial, and sex-based heterogeneity in recurrence risk— an gap that limits equitable and personalized care. To evaluate the associations of demographic factors, obesity, and adenoma features with recurrence risk over time in a large longitudinal surveillance cohort.

**Methods:** This retrospective cohort study included 59,667 adults who underwent their first colonoscopic polypectomy between January 1990 and July 2024 at a tertiary medical center. Median follow-up was 4 years. Demographic variables included race and ethnicity (non-Hispanic White [NHW], non-Hispanic Black [NHB], Hispanic, Asian or Pacific Islander [API]), sex, obesity (BMI >30), family history of colorectal cancer (CRC) or polyps, and age at adenoma onset (<50 vs ≥50 years). Adenoma features included histology, size, number, and dysplasia. The primary outcome was recurrence-free survival, defined as time from initial polypectomy to histologically confirmed recurrence. Cox proportional hazards models estimated associations adjusted for confounders, with stratified analyses over 5-, 10-, and >10-year follow-up intervals.

**Findings:** Among 59,667 patients, 17,596 (29.5%) experienced recurrence within 5 years, revealing substantial temporal heterogeneity. Early recurrence was associated with male sex (adjusted hazard ratio [aHR], 1.10; 95% CI, 1.06–1.14), obesity (aHR, 1.18; 95% CI, 1.13–1.23), early-onset adenomas (aHR, 1.17; 95% CI, 1.11–1.23), and family history of CRC (aHR, 1.24; 95% CI, 1.18–1.31). Compared with NHW patients, NHB individuals had lower early recurrence risk (aHR, 0.89; 95% CI, 0.83–0.96) but higher late recurrence (>10 years; aHR, 1.26; 95% CI, 1.06–1.50). API patients had a similar shift, with lower early risk (aHR, 0.80; 95% CI, 0.67– 0.96) and elevated mid-term risk (5–10 years; aHR, 1.40; 95% CI, 1.08–1.81). High-grade dysplasia (aHR 2.86; 95% CI, 2.54–3.22) and villous histology (aHR 2.55; 95% CI, 2.31–2.81showed the largest effect sizes for early recurrence. Females had stronger associations with tubulovillous histology, mixed adenomas, and large lesions.

**Interpretation:** Temporal, demographic, and histologic differences in adenoma recurrence highlight the need for surveillance strategies that incorporate population- and time-specific risk profiles to enhance colorectal cancer prevention.

**Funding:** This work was supported by the National Cancer Institute (Grant No. R37CA227130 to Xingyi Guo).

**Research in context:** *Evidence before this study:* We conducted a PubMed search for publications dated before June 2024 using combinations of keywords such as “colonoscopic polypectomy,” “Demographic and Clinicopathologic Risk Factors,” “Vannderbilt,” and “electronic health records.” We found no studies that comprehensively evaluated the associations of demographic characteristics, obesity, and adenoma features with recurrence risk over time in a large, longitudinal surveillance cohort.

*Added Value of This Study:* Using a longitudinal cohort of 59,667 patients, our study reveals substantial temporal heterogeneity in adenoma recurrence. Non-Hispanic Black and Asian or Pacific Islander individuals exhibited a lower risk of recurrence within the first 5 years but experienced increased risk at 5–10 and >10 years post-polypectomy. Females showed heightened early recurrence risk, particularly when initial adenomas were tubulovillous, mixed-type, or large. Early recurrence was predominantly driven by high-grade dysplasia, high-risk adenomas, villous or tubulovillous histology, and multiplicity.

*Implications of All the Available Evidence:* These findings highlight the critical need to recognize and address temporal, racial, and sex-specific heterogeneity in adenoma recurrence risk. The observed variability in histopathologic and demographic factors over time underscores the importance of personalized, adaptive surveillance strategies to reduce adenoma recurrence and enhance colorectal cancer prevention.

## Introduction

Colorectal cancer (CRC) persists as a leading cause of cancer-related mortality globally, with adenomatous polyps representing the primary precursor lesions for 60–90% of cases ^1,2^. Current post-polypectomy surveillance guidelines, such as those from the U.S. Multi-Society Task Force (USMSTF) and the European Society of Gastrointestinal Endoscopy (ESGE), stratify recurrence risk primarily based on polyp characteristics, including size, histology, and multiplicity ^1–3^. Recent updates to these guidelines, including the USMSTF recommendations of 7–10 years for 1–2 nonadvanced adenomas (NAAs) and the 2020 ESGE’s omission of surveillance for 1–4 NAAs, reflect evolving evidence but reveal persistent inconsistencies. For instance, the ESGE and British Society of Gastroenterology/Association of Coloproctology of Great Britain and Ireland/Public Health England (BSG/ACPGBI/PHE) guidelines recommend a return to routine screening for 1–4 NAAs (tubular adenomas <10 mm with low-grade dysplasia), whereas the USMSTF advocates surveillance intervals ranging from 3–10 years, depending on adenoma number ^1,3,4^.

These discrepancies underscore a broader limitation: guidelines universally neglect demographic variables—such as race, sex, family history of polyps, and family history of CRC, and age—and modifiable risk factors like obesity, potentially perpetuating inequities in colorectal adenoma recurrence and CRC prevention ^5–7^. Of note, racial disparities in CRC outcomes are well-documented, with non-Hispanic Black (NHB) individuals experiencing higher CRC incidence and mortality compared to White populations; however, limited studies have explored the association between demographic disparities and adenoma recurrence ^8,9^. Despite differential recurrence patterns across ethnic groups, current surveillance protocols lack race-specific risk stratification, while central obesity (waist-to-hip ratio) independently increases CRC risk by 15–18% per standard deviation, surpassing BMI’s effect ^6,7^.

Histopathological features, such as high-grade dysplasia and villous architecture, are recognized as having significant effect sizes in their association with colorectal adenoma recurrence ^9–11^. However, their prioritization varies across current clinical guidelines, contributing to inconsistent surveillance recommendations. USMSTF classifies villous histology or high-grade dysplasia as high-risk for 3-year surveillance, but ESGE prioritizes size and dysplasia, potentially underestimating villous histology’s long-term impact, with neither addressing racial or sex-specific recurrence variations ^9,12^. Compounding these issues, long-term surveillance data from racially and ethnically diverse populations remain sparse, restricting insights into how demographic and histopathological factors interact to influence post-polypectomy outcomes over time ^9,13,14^. Investigating recurrent colorectal adenoma demands longitudinal follow-up with serial colonoscopies to confirm polyp recurrence and polyp-free status in controls.

Current literature provides limited insight into temporal recurrence dynamics—namely, how risk patterns evolve over extended follow-up—thus constraining the optimization of long-term surveillance strategies ^1,2,15,16^. Although guidelines assume constant risk, obesity and male sex drive recurrence within 3–5 years post-polypectomy, while villous histology, lesion multiplicity, and high-grade dysplasia show persistent or re-emerging risk after 10 years, necessitating temporally adaptive surveillance frameworks^17–19^.

To overcome these barriers, we analyzed Vanderbilt electronic health records (EHRs), covering diagnoses, procedures, and pathology reports from over 3.5 million patients, using a dataset of 59,667 Polypectomy Cohort individuals to clarify post-polypectomy surveillance determinants, focusing on obesity, demographic disparities, and polyp characteristics, thereby providing a foundation for refining polyp-centric risk models and establishing targeted surveillance strategies for high-risk populations.

## Methods

### Study Design and Setting

The Vanderbilt Polypectomy Cohort was recently established to investigate adenoma recurrence patterns across demographic subgroups using clinical data from Vanderbilt University Medical Center (VUMC) spanning January 1990 through July 2024. To ensure consistency and accuracy despite changes in clinical practice, diagnostic standards, and coding systems over the years—including the transition from ICD-9 to ICD-10—harmonized data extraction protocols were meticulously implemented ^20^.

### Data Source

Data was extracted from VUMC EHR system (Epic Systems Corporation’s eStar), encompassing colonoscopy reports, pathology findings, demographics, and comorbidities. To ensure robustness, a multistep validation process was implemented. First, polyp cases and procedures were identified using standardized diagnostic and procedural codes (ICD-9, ICD-10, CPT, HCPCS) to address discrepancies across coding systems. Second, all polyp diagnoses were cross verified against original histopathology reports to confirm neoplastic status, histology, and dysplasia grade. Third, a secure institutionally managed large language model (LLM) was deployed to extract adenoma characteristics—including polyp histology, size, and number—from unstructured clinical notes and pathology reports, with a verification rate > 95% based on our comparison between procedural codes and LLM from randomly selected 100 patient records ^21^.

### Vanderbilt Polypectomy Cohort Identification

The study population comprised adults aged ≥18 years with histopathologically confirmed colorectal adenoma who underwent polypectomy at VUMC. Exclusion criteria were prior CRC and other cancers, non-neoplastic polyps, colectomy, or incomplete colonoscopies. Colorectal adenomas were categorized as high-risk if they met one or more criteria: ≥3 number, size ≥10 mm, villous/tubulovillous histology, or high-grade dysplasia. Low-risk adenomas were defined as <3 adenomas, <10 mm in size, and low-grade dysplasia. A multidisciplinary team (U.A.A., Q.S., Z.Y., X.G.) independently reviewed cohort eligibility to ensure adherence to clinical and histopathologic standards. Additional details on data extraction and variable definitions are provided in the Supplemental Methods.

### Follow-Up and Censoring

Patients were followed from the index polypectomy until the earliest occurrence of adenoma recurrence, death, loss to follow-up, or the study’s end in July 2024. Longitudinal recurrence patterns were primarily evaluated using a 5-year follow-up interval, with events after this period censored, while additional intervals of <5 years, 5-10 years, and >10 years were assessed to explore trends, as detailed in the supplementary files (eTable 1, 2), censoring events after 5 years for the <5-year analysis, after 10 years for the 5-10-year analysis, and analyzing events beyond 10 years for the >10-year interval. Patients undergoing colectomy (complete or partial) or lost to follow-up were censored at their last documented clinical contact.

The primary outcome, recurrence-free survival, was defined as the time from initial polypectomy to histologically confirmed recurrence on subsequent colonoscopy (≥6 months post-procedure). This was analyzed both as a binary endpoint and as time-to-event data to estimate recurrence-free survival. Recurrence timing was evaluated across pre-defined intervals (<5, 5–10, and >10 years), and incorporated into multivariable Cox regression models to assess associations with demographic, clinical, and histopathological factors. Censoring rules and follow-up windows were applied as detailed in the corresponding analytical framework.

### Statistical Analysis

Baseline characteristics were summarized using descriptive statistics, with categorical variables reported as numbers and percentages (%). Cox proportional hazard models were used to estimate hazard ratios (HRs) and 95% confidence intervals (CIs), adjusting for potential confounders including race/ethnicity, sex, obesity, family history of polyp, family history of CRC, NSAID, Aspirin use (during the post-polypectomy period) and polyp characteristics (number, size, and type), and baseline adenoma risk (high vs low). Recurrence patterns were primarily assessed using a 5-year follow-up interval, with extended intervals up to and beyond 10 years evaluated to explore long-term trends, censoring events after each interval as appropriate. The proportional hazards assumption was verified using Schoenfeld residuals, and survival curves were generated via the Kaplan-Meier method to compare recurrence-free probabilities across demographic and clinical subgroups. Stratified analyses were conducted by race/ethnicity, sex, recurrence status, and follow-up interval (< 5 Years, 5-10 years and >10-year windows). Statistical significance was defined as a two-tailed *P* < 0.05.

## Results

The Vanderbilt Polypectomy Cohort included 59,667 patients who underwent colon polypectomy, with a median follow-up of 4 years (Figure 1). The cohort consisted of 29,401 females (49.3%) and 30,266 males (50.7%), with a racial/ethnic distribution of 52,042 (NHW, 87.2%), 5,972 (NHB, 10.0%), 1,007 (API, 1.7%), and 646 Hispanic (1.1%) individuals. Early-onset adenoma was identified in 11,018 patients (18.5%), while late-onset adenoma was observed in 48,649 patients (81.5%). Adenoma characteristics varied significantly by sex and race/ethnicity. Males exhibited higher prevalence of tubulovillous (4.3% vs. 3.6% in females) and mixed adenomas (27.9% vs. 27.2%), while females had more serrated (5.9% vs. 3.4%) and villous adenomas (3.5% vs. 2.2%; *P* = 1.4 × 10□□). Among racial groups, NHW patients showed elevated mixed (27.7%) and serrated adenoma rates (4.8%), NHB had higher tubular (36.3%) and tubulovillous adenomas (5.4%), and API had the highest tubular adenoma prevalence (40.0%; *P* = 1.6 × 10□□). Early-onset adenomas were more common in Hispanic (30.5%) and API (20.3%) than NHW patients (18.2%; *P* = 2.6 × 10□¹□). Family history of CRC was more frequent in females (15.7% vs. 11.9% in males; *P* = 4.9 × 10□□²) and NHW (14.0%; *P* = 1.7 × 10□□), while family history of polyps was higher in females (3.2% vs. 2.1%; *P* = 2.5 × 10□¹□) and NHW (2.8%; *P* = 5.5 × 10□¹□). High-risk adenomas were slightly more prevalent in males (25.3% vs. 23.8%; *P* = 5.6 × 10 ) (Table 1).

**Figure 1:**
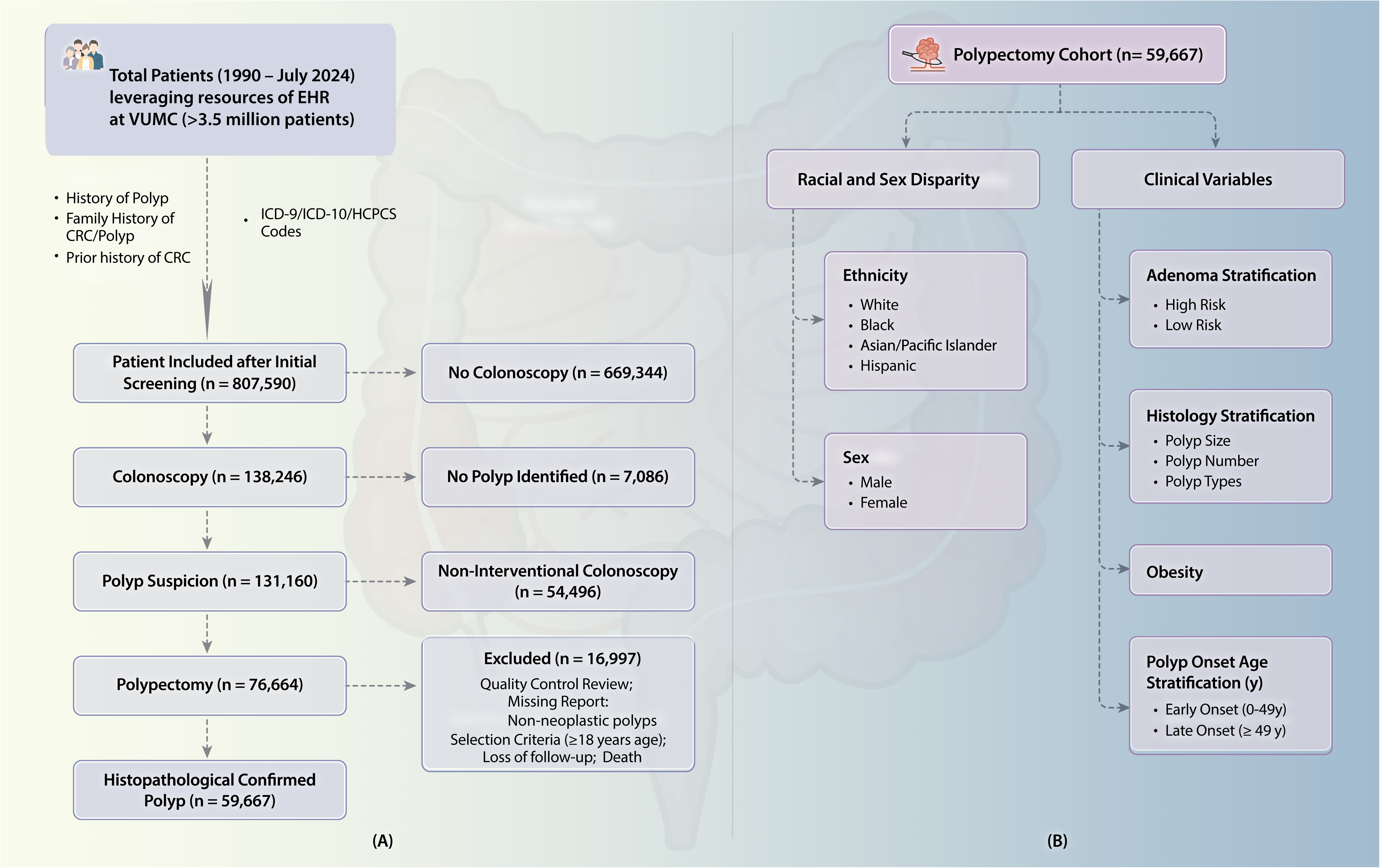
Patient Selection and Cohort Characteristics for the Vanderbilt Polypectomy Cohort. **(A):** Flowchart illustrates the inclusion and exclusion criteria for patients undergoing colonoscopy and polypectomy from 1990 to July 2024, resulting in a final cohort of 59,667 patients. (B): Stratification of the polypectomy cohort by demographic factors (ethnicity and sex) and clinical variables (adenoma risk, histology, obesity, and age at polyp onset).

**Table 1:**
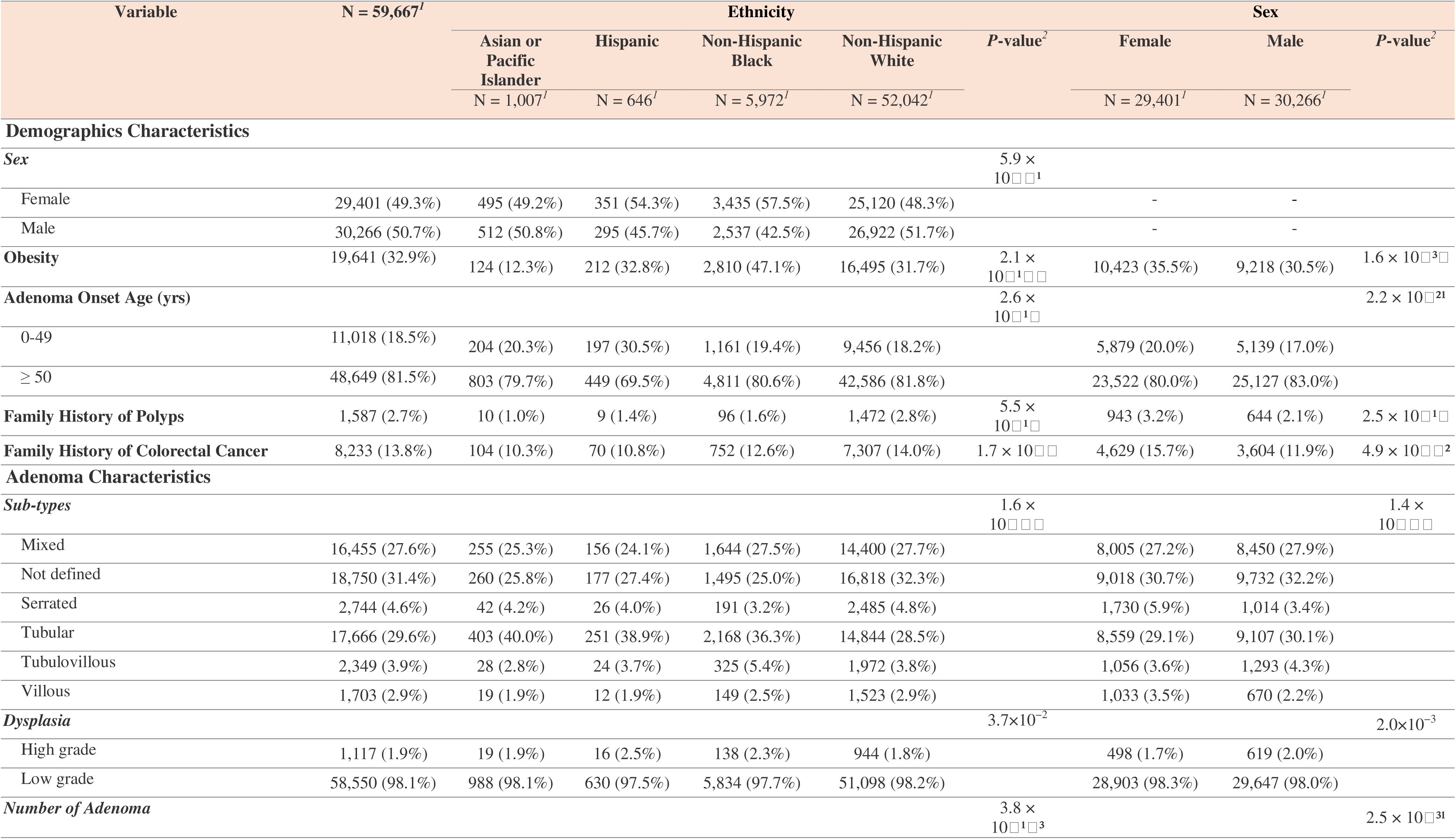
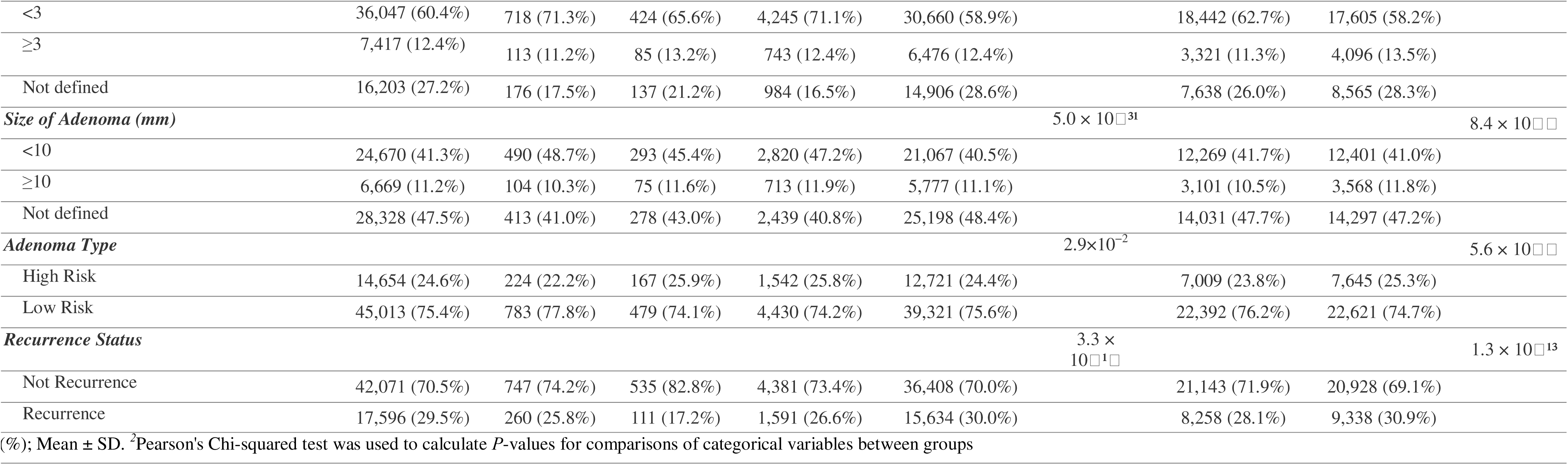
Baseline Characteristics of the Study Population Stratified by Race/Ethnicity and Sex.

### Recurrence Rates and Demographic-Clinical Associations

Adenoma recurrence occurred in 17,596 patients (29.5%) within 5 years, with significant associations observed across demographic and clinical variables (Table 2). Males showed a higher recurrence rate than females (30.9% vs. 28.1%; adjusted hazard ratio [aHR] 1.1, 95% CI 1.06–1.14; *P* = 3.46 × 10□□). By race/ethnicity, NHW patients had a 30.0% recurrence rate, compared to 26.6% in NHB (aHR 0.89, 95% CI 0.83–0.96; *P* = 1.97 × 10□³), 25.8% in API (aHR 0.8, 95% CI 0.67–0.96; *P* = 1.36 × 10□□²), and 17.2% in Hispanic patients (aHR 0.89, 95% CI 0.70–1.14; *P* = 3.63 × 10□¹). Obesity, present in 19,641 patients (32.9%), was associated with elevated recurrence risk (32.3% vs. 67.7% non-obese; aHR 1.18, 95% CI 1.13–1.23; *P* = 1.84 × 10□¹□). Patients with early-onset adenomas had a 26.4% recurrence rate, demonstrating a higher risk than those with late-onset (30.2%; aHR 1.17, 95% CI 1.11–1.23; *P* = 6.77 × 10□□). Family history of CRC exhibited a strong association with recurrence (38.7%; aHR 1.24, 95% CI 1.18– 1.31; *P* = 4.44 × 10□¹□), particularly in females and NHW patients. Family history of polyps showed a weaker association (38.5%; aHR 1.04, 95% CI 0.93–1.17; *P* = 4.64 × 10□□¹).

**Table 2:** Results of Adenoma Recurrence associated with Demographic Variables for 5-Year Survival Analysis.

| Variable | Not Recurrence | Recurrence | P-value<sup>2</sup> | Adjusted Hazard Ratio* |  |  |
| --- | --- | --- | --- | --- | --- | --- |
|  | N = 42,071<sup>1</sup> | N = 17,596<sup>1</sup> |  | HR<sup>3</sup> | 95% CI<sup>4</sup> | P-value |
| Demographic Characteristics |  |  |  |  |  |  |
| Sex<sup>5</sup> |  |  | 1.3 × 10<sup>−13</sup> |  |  |  |
| Female | 21,143 (71.9%) | 8,258 (28.1%) |  | — | — |  |
| Male | 20,928 (69.1%) | 9,338 (30.9%) |  | 1.1 | 1.06, 1.14 | 3.46 × 10<sup>−11</sup> |
| Ethnicity<sup>6</sup> |  |  | 3.3 × 10<sup>−11</sup> |  |  |  |
| Non-Hispanic White | 36,408 (70.0%) | 15,634 (30.0%) |  | — | — |  |
| Asian or Pacific Islander | 747 (74.2%) | 260 (25.8%) |  | 0.8 | 0.67, 0.96 | 1.36 × 10<sup>−2</sup> |
| Hispanic | 535 (82.8%) | 111 (17.2%) |  | 0.89 | 0.70, 1.14 | 3.63 × 10<sup>−1</sup> |
| Non-Hispanic Black | 4,381 (73.4%) | 1,591 (26.6%) |  | 0.89 | 0.83, 0.96 | 1.97 × 10<sup>−3</sup> |
| Obesity<sup>7</sup> | 13,302 (67.7%) | 6,339 (32.3%) | 1.5 × 10<sup>−21</sup> | 1.18 | 1.13, 1.23 | 1.84 × 10<sup>−11</sup> |
| Adenoma Onset Age (yr)<sup>8</sup> |  |  | 8.7 × 10<sup>−11</sup> |  |  |  |
| 0-49 | 8,104 (73.6%) | 2,914 (26.4%) |  | 1.17 | 1.11, 1.23 | 6.77 × 10<sup>−11</sup> |
| ≥ 50 | 33,967 (69.8%) | 14,682 (30.2%) |  | — | — |  |
| Family History of Polyps<sup>9</sup> | 976 (61.5%) | 611 (38.5%) | 1.5 × 10<sup>−11</sup> | 1.04 | 0.93, 1.17 | 4.64 × 10<sup>−1</sup> |
| Family History of Colorectal Cancer<sup>10</sup> | 5,044 (61.3%) | 3,189 (38.7%) | 2.4 × 10<sup>−11</sup> | 1.24 | 1.18, 1.31 | 4.44 × 10<sup>−11</sup> |

<sup>1</sup>n (%); <sup>2</sup>Pearson's Chi-squared test; <sup>3</sup>HR = Hazard Ratio, <sup>4</sup>CI = Confidence Interval.
Model adjustments: <sup>5</sup>adjusted for ethnicity; stratified by age groups; <sup>6</sup>adjusted for sex; stratified by age group; <sup>7</sup>adjusted for sex and ethnicity; stratified by age group; <sup>8</sup>adjusted for sex, ethnicity, use of Non-steroidal anti-inflammatory drugs (NSAIDs), use of Aspirin, family history of colorectal cancer, and family history of polyps; stratified by obesity and age group; <sup>9</sup>adjusted for sex, ethnicity, use of NSAIDs, use of Aspirin, and family history of colorectal cancer; stratified by obesity, Adenoma Onset Stratification, and age group; <sup>10</sup>adjusted for sex, ethnicity, use of NSAIDs, use of Aspirin, and family history of polyps; stratified by obesity, Adenoma Onset Stratification, and age group.

### Adenoma Characteristics and Recurrence

Adenoma characteristics demonstrated significant associations with recurrence, with histopathological features exhibiting the largest effect sizes (Table 3). Villous histology had the highest recurrence rate (57.7%; aHR 2.55, 95% CI 2.31–2.81; *P* = 4.55 × 10□□□), followed by tubulovillous (32.6%; aHR 2.18, 95% CI 1.99–2.39; *P* = 1.58 × 10□□¹) and mixed adenomas (31.2%; aHR 1.37, 95% CI 1.30–1.45; *P* = 6.99 × 10□□³²), compared to tubular adenomas (28.2%). Serrated adenomas showed a moderate association (28.1%; aHR 1.3, 95% CI 1.18– 1.44; *P* = 1.10 × 10□□). High-grade dysplasia was strongly associated with recurrence (30.9%; aHR 2.86, 95% CI 2.54–3.22; *P* = 4.13 × 10□□□) relative to low-grade dysplasia (29.5%). Multiple adenomas (≥3) had a recurrence rate of 33.5% (aHR 1.69, 95% CI 1.60–1.79; *P* = 5.95 × 10□□□), higher than fewer than 3 (28.3%). Adenomas ≥10 mm showed increased risk (30.8%; aHR 1.63, 95% CI 1.53–1.73; *P* = 5.72 × 10 ), though with a smaller effect size than histopathological features. High-risk adenomas, observed in 24.6% of patients, had a 34.5% recurrence rate (aHR 2.79, 95% CI 2.68–2.91; *P* = 2.22 × 10□¹□), significantly exceeding low-risk adenomas (27.9%). Adjusted survival curves for polyp type, dysplasia grade, number, and size over 5 years are illustrated in Figure 2.

**Figure 2:**
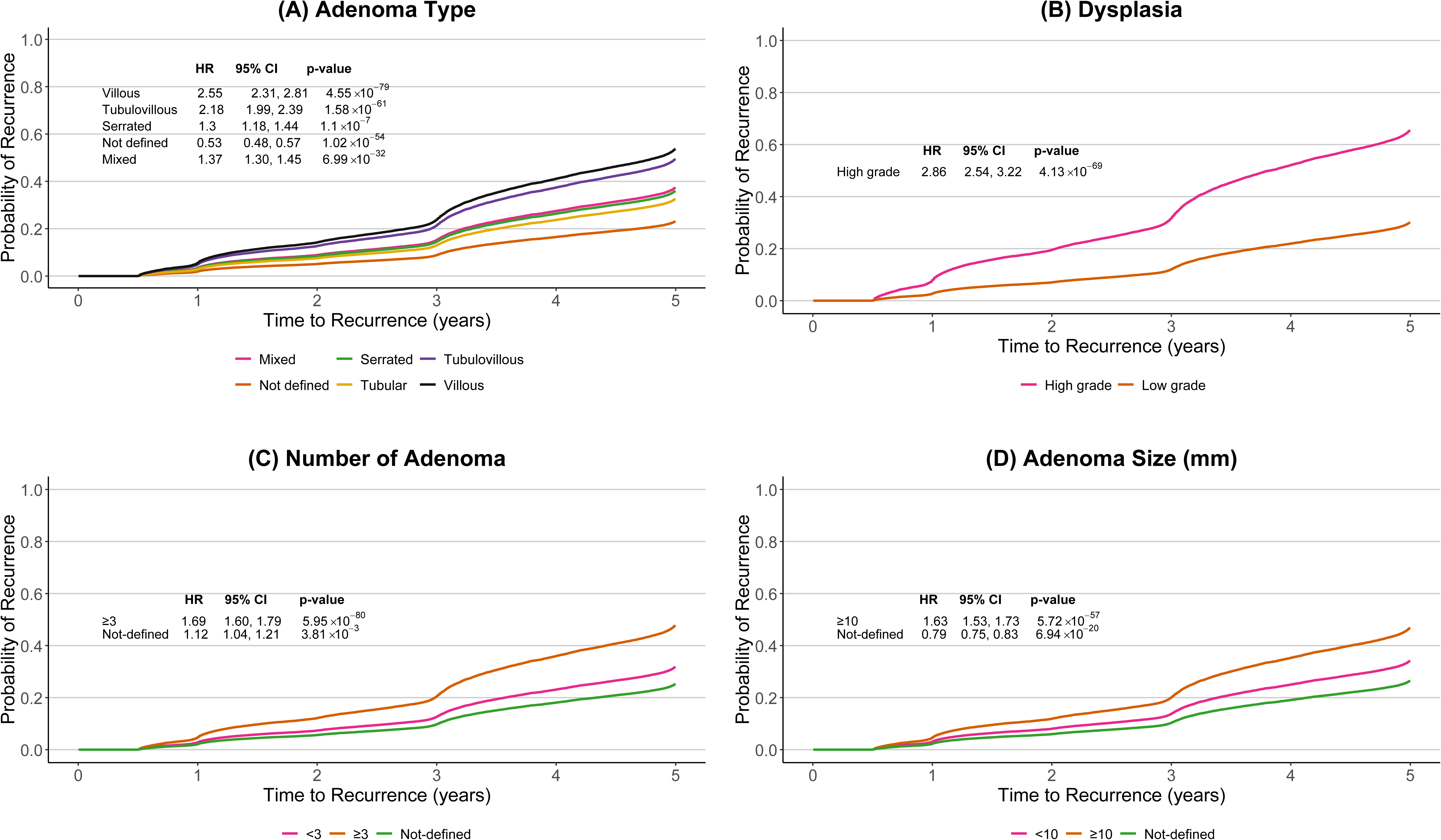
Adjusted Survival Curves for Time to Adenoma Recurrence After Polypectomy in the Vanderbilt Polypectomy Cohort. (A) Survival by polyp type. (B) Survival by dysplasia grade. (C) Survival by number of polyps. (D) Survival by polyp size (mm). Survival curves are derived from Cox proportional hazards models adjusted for ethnicity, sex, polyp onset age, obesity, use of NSAIDs, use of aspirin, family history of colorectal cancer, and family history of polyps, plotted for a 5-year follow-up period. *P*-values are obtained from the adjusted Cox proportional hazards models. aHR, hazard ratio; CI, confidence interval.

**Table 3:**
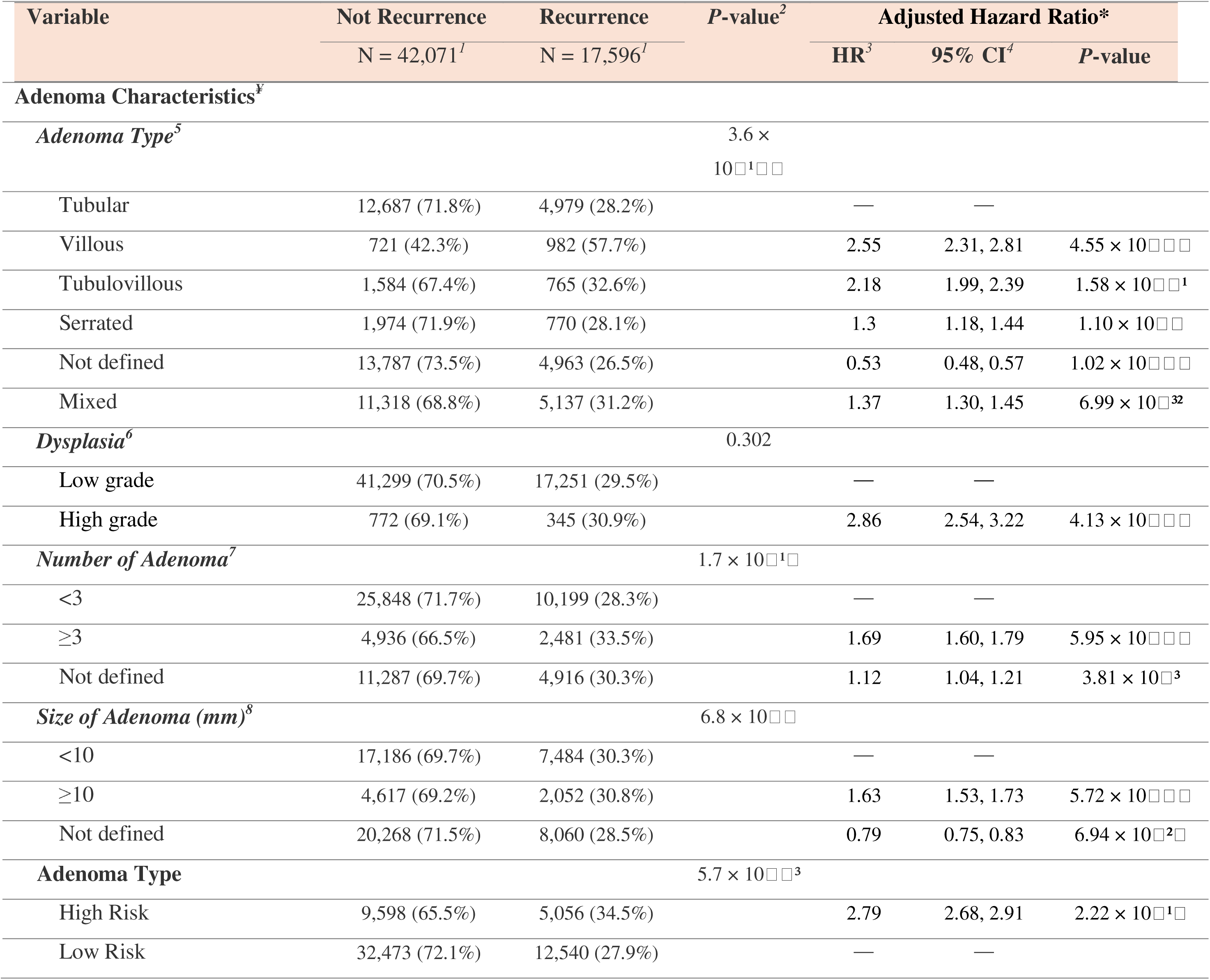
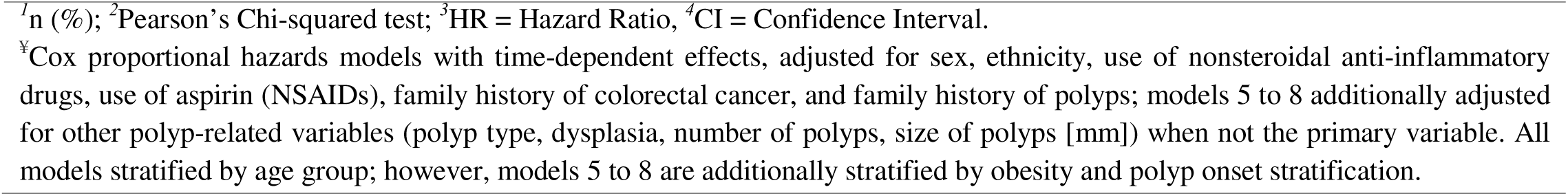
Results of Adenoma Characteristics Associated with Recurrence for 5-Year Survival Analysis.

### Long-Term Recurrence Patterns and Temporal Heterogeneity

Colorectal adenoma recurrence demonstrated pronounced temporal heterogeneity over the 25-year surveillance period (eTables 1–2). Among racial groups, NHB patients exhibited a temporal shift from reduced early recurrence risk (<5 years; aHR 0.89, 95% CI 0.83–0.96; *P* = 1.97 × 10□³) to elevated late-phase hazard (>10 years; aHR 1.26, 95% CI 1.06–1.50; *P* = 9.22 × 10□□³). Similarly, API patients experienced early protection (aHR 0.80, 95% CI 0.67–0.96; *P* = 1.36 × 10□²) followed by heightened mid-term recurrence risk (5–10 years; aHR 1.40, 95% CI 1.08–1.81; *P* = 1.04 × 10□²). Temporal recurrence risk was most strongly influenced by histopathologic features during the early phase, including high-grade dysplasia (aHR 2.86, 95% CI 2.54–3.22; *P* = 4.13 × 10□□), high-risk adenomas (aHR 2.79, 95% CI 2.68–2.91; *P* = 2.22 × 10□¹□), and villous histology (aHR 2.55, 95% CI 2.31–2.81; *P* = 4.55 × 10□□□), with attenuation of these associations over long-term follow-up. Tubulovillous histology (aHR 2.18, 95% CI 1.99–2.39; *P* = 1.58 × 10□□¹) also conferred elevated early recurrence hazard, reinforcing the importance of temporally adaptive, pathology-specific surveillance strategies.

### Heterogeneity of Effects by Race and Sex

Five-year stratified analyses revealed significant sex- and race-specific heterogeneity in recurrence risk (eTables 3–4), reinforcing the rationale for personalized surveillance. Females exhibited significantly greater early recurrence hazards for tubulovillous histology (aHR 2.21, 95% CI 1.93–2.53; *P* = 2.67 × 10□³□), mixed adenomas (aHR 1.43, 95% CI 1.32–1.55; *P* = 3.19 × 10□¹□), and large lesions (≥10 mm; aHR 1.60, 95% CI 1.46–1.74; *P* = 2.77 × 10□²□), with stronger associations than those observed in males relative to tubular or smaller (<10 mm) lesions. Among racial groups, NHB patients exhibited significantly higher recurrence risk for multiple adenomas (aHR 1.67, 95% CI 1.42–1.98; *P* = 1.85 × 10□□), indicating a greater burden of lesion multiplicity. These findings underscore the imperative for demographically stratified surveillance paradigms to optimize post-polypectomy care.

## Discussion

Current Vanderbilt Polypectomy Cohort, encompassing 59,667 patients who underwent polypectomy, provides critical insights into the demographic and clinicopathological factors driving colorectal adenoma recurrence and long-term outcomes. This study, with its 25-year follow-up, reveals distinct recurrence patterns across sex, age, race, and histopathological subtypes, highlighting opportunities to refine current post-polypectomy surveillance guidelines (e.g., USMSTF and ESGE) through personalized, data-driven risk stratification approaches that enhance precision and equity in CRC prevention.

Males exhibited higher early recurrence risk, driven by tubulovillous and mixed adenomas, consistent with studies linking male sex to advanced neoplasia ^22–25^. Females showed increased risk with villous architecture and larger adenoma, independent of adenoma burden, suggesting a sex-histology-size interaction. This challenges USMSTF’s uniform 3–10-year surveillance intervals^9^, which overlook sex as a risk modifier. Shorter intervals for females with villous morphology could enhance precision prevention.

Recurrence patterns varied by race/ethnicity. NHB patients displayed delayed but elevated recurrence, contrasting with early peaks in API and Hispanic populations. Despite higher tubulovillous adenoma burden, NHB patients had lower early recurrence, possibly due to lower adenoma detection rates and delayed follow-up, exacerbating disparities ^26–28,45,49^. Current guidelines (USMSTF, ESGE, BSG/ACPGBI/PHE) inadequately address this heterogeneity, with ESGE omitting surveillance for villous adenomas <10 mm ^2,15,16,36,37^. Race-stratified surveillance—extended for NHB patients and tighter early intervals for those with familial CRC risk—could bridge equity gaps. Prior studies report inconsistent racial differences in advanced neoplasia ^29–31^, suggesting adenoma characteristics and surveillance intensity influence outcomes. Structural inequities, including unequal healthcare access and lower screening rates among NHB patients ^29–31,45,49^, necessitate standardized care and prospective multi-institutional studies.

Early-onset adenomas, prevalent among Hispanic and API patients, were associated with higher early recurrence risk, aligning with rising early-onset CRC trends ^32,33^. Despite USMSTF’s 2022 recommendation for screening at age 45 ^34,35^, guidelines do not account for age at adenoma detection as a recurrence modifier. Family history of CRC was associated with early recurrence risk (aHR 1.24), while polyp family history showed a stronger effect size for recurrence at 5–10 years, indicating temporally distinct trajectories ^36–40^. Integrating onset age and familial subtype into risk models could refine surveillance intervals. Obesity (32.9% prevalence) was associated with recurrence (aHR 1.18), with stronger effects in females, possibly due to visceral adiposity and endocrine signaling ^41–43^.

Villous architecture and high-grade dysplasia exhibited the largest effect sizes for recurrence, surpassing adenoma number (≥3: aHR 1.69) and size (≥10 mm: aHR 1.63). Villous adenomas conferred sustained risk beyond 10 years, while high-grade dysplasia elevated early risk. Females with villous features or larger adenomas faced higher recurrence, supporting sex-histology-size-specific stratification. USMSTF flags villous histology as high-risk, advocating 3-year surveillance ^35,44^, but ESGE’s omission of villous features ^2^ may underestimate risk. High-risk adenomas exhibited persistent hazard, with late-phase resurgence (>10 years), underscoring the need for prolonged, individualized follow-up prioritizing histologic severity.

Adenoma recurrence risk is heterogeneous and dynamic, inadequately captured by static guideline frameworks. Time-stratified Cox models revealed evolving risk trajectories: obesity’s influence dissipated after 5 years, early-onset lesions shifted from excess to deficit risk, and NHB patients transitioned from early protection to late excess risk. High-risk adenomas showed a late-phase resurgence (>10 years), unlike the time-limited effects of dysplasia and obesity. Villous morphology maintained a persistent hazard, even in diminutive adenomas, challenging ESGE’s exclusion of villous features <10 mm ^2^. A recent cohort noted higher adenoma burden in NHB at older ages, suggesting biology-care interactions ^45^, while a study found no race effect with uniform colonoscopy quality ^31^. These findings reconcile discordant results, emphasizing dynamic, race-specific hazards. The 2020 USMSTF lengthened surveillance for low-risk adenomas but retained uniform high-risk intervals^35^, and ESGE’s simplified high-risk definitions ^2,55^ overlook sustained villous risk. Integrating temporal coefficients and heterogeneity metrics could align surveillance with evolving patient risk, enhancing precision and equity in CRC prevention.

This study’s cohort of 59,667 patients, followed up to 25 years, robustly examines colorectal adenoma recurrence across diverse subgroups using validated natural language processing for histologic and clinical feature extraction, enabling stratified analyses by sex, race/ethnicity, onset age, and adenoma subtype. Time-dependent Cox models reveal dynamic recurrence trajectories over <5, 5–10, and >10-year intervals, uncovering late-phase risks often missed, with real-world EHR data enhancing generalizability. However, this single-center study, predominantly non-Hispanic White, may not fully generalize to diverse populations, and its retrospective design risks selection bias and confounding from unmeasured factors (e.g., diet, smoking, socioeconomic status, healthcare access, colonoscopy quality). Variable surveillance frequency across groups may affect recurrence detection, and reliance on VUMC network colonoscopies risks misclassification if patients were surveilled elsewhere.

## Conclusion

In this cohort of 59,667 patients, villous histology, high-grade dysplasia, and adenoma multiplicity emerged as dominant recurrence drivers, though their hazards varied by time and population. Risk peaked early among males, obese patients, and those with early-onset lesions, but shifted later among non-Hispanic Black individuals. Notably, villous features conferred a stronger recurrence hazard in females. Incorporating these temporally dynamic, demography-specific risks into surveillance algorithms may enhance precision, reduce disparities, and help current guidelines to enhance precision and equity in post-polypectomy surveillance.

## Data Availability

All data produced in the present study are available upon reasonable request to the authors.

## Contributors

X.G. and Y.Z. conceived and designed the study. U.A.A., Q.S., W.W., Z.Y., and X.G. collected the data and developed the analytical plan. U.A.A., Q.S., Z.Y., and X.G. accessed, verified, and processed the raw cohort data used in this study. U.A.A., together with Q.S., W.W., Z.Y., and X.G., drafted the manuscript with input from all co-authors. All authors contributed to data interpretation, manuscript revision, and approved the final version for submission. All authors had full access to all the data in the study and shared final responsibility for the decision to submit the manuscript for publication.

## Declaration of interest

None of the authors have any conflicts of interest to disclose.

## Data sharing

The datasets used and analyzed in the current study are available from the corresponding authors upon reasonable request.

## Notes

### Competing Interest Statement

The authors have declared no competing interest.

### Funding Statement

This study did not receive any funding.

### Author Declarations

This study was reviewed and exempted from human subjects' research by the Institutional Review Board at Vanderbilt University Medical Center (IRB #250158). Patient confidentiality was maintained through data de-identification, and all procedures adhered to relevant ethical standards. Data was stored and analyzed in a secure, encrypted, access-controlled database.

### Summary of Updates

Cox proportional hazards models estimated associations adjusted for confounders, with stratified analyses over 5-, 10-, and >10-year follow-up intervals. The results have been updated based on this new analysis.

