## Supplementary_File for "Demographic and Clinicopathologic Risk Factors for Colorectal Adenoma Recurrence: A Large-Scale Surveillance Cohort Study of 59,667 Adults"

**Author Affiliations:**

### Supplemental Methodology

#### Data Extraction and Variable Definitions

Variables were extracted and categorized as follows: Demographic factors included age (stratified as ≤50, 51–75, ≥76 years), sex (male/female), self-reported race/ethnicity (non-Hispanic White [NHW], NHB, Hispanic, Asian/Pacific Islander [API]), family history of polyp, and family history of CRC. Clinical variables included obesity (BMI ⩾30 kg/m$^2$ , defined via ICD-9/ICD-10 codes), medication use (e.g., aspirin, NSAIDs) and screening colonoscopies (procedures without biopsy). Adenoma characteristics encompassed histological subtype (tubular, tubulovillous, villous, serrated, mixed), number (<3 vs. ≥3), size (<10 mm vs. ≥10 mm) and dysplasia grade (low/high). Mixed adenoma was defined as the presence of more than one histologic polyp subtype (e.g., tubular, villous, serrated) within a single patient at the time of index colonoscopy. For analysis of polyp onset age, patients were categorized into two groups: early onset (0–49 years), and late onset (⩾50 years).

Furthermore, a secure LLM managed by VUMC was used to extract polyp characteristics—including type, number, and size—from unstructured histopathology reports and clinical notes. The LLM utilized case-insensitive keyword searches to identify relevant terms such as *polypectomy*, *adenoma*, *tubul*, *villous*, *sessile*, *plastic*, and *serrated* within clinical documentation (Supplement 1). Variables with incomplete or unextractable data (e.g., missing histology, size, or multiplicity) were systematically labeled as *Not Defined* to preserve data integrity and transparency.

### Ethical Considerations

This study was reviewed and exempted from human subjects' research by the Institutional Review Board at Vanderbilt University Medical Center (IRB #250158). Patient confidentiality was maintained through data de-identification, and all procedures adhered to relevant ethical standards. Data was stored and analyzed in a secure, encrypted, access-controlled database.

### Table of Contents

**eTable1.** Adjusted Hazard Ratios for Adenoma Recurrence by Demographic and Clinical Variables Across Time Periods……………….4

**eTable 2.** Adjusted Hazard Ratios for Adenoma Recurrence by Adenoma Characteristics Across Time Periods……………………………5

**eTable 3.** Adjusted Hazard Ratios for Adenoma Recurrence by Adenoma Characteristics Across Ethnicities for 5 year follow-up…………...6

**eTable 4:** Adjusted Hazard Ratios for Adenoma Recurrence by Adenoma Characteristics Across Sex for 5 year follow-up…………………..7

**Supplement 1**. Pathology Report Extraction……………………………………………………………………………….8

**eTable 1.** Adjusted Hazard Ratios for Adenoma Recurrence by Demographic and Clinical Variables Across Time Periods.

| Variables | < 5 Years | | | 5-10 Years | | | > 10 Years | | | Heterogeneity | | |
|---|---|---|---|---|---|---|---|---|---|---|---|---|
| | **HR** | **95% CI** | ***P*-Value** | **HR** | **95% CI** | ***P*-Value** | **HR** | **95% CI** | ***P*-Value** | **Cochran's Q** | ***P*-value** | **$I^2$ (%)** |
| **Demographic Characteristics** | | | | | | | | | | | | |
| ***Sex*[1]** | | | | | | | | | | | | |
| Female | - | - | - | - | - | - | - | - | - | - | - | - |
| Male | 1.1 | 1.06, 1.14 | $3.46 \times 10^{-6}$ | 1.06 | 0.99, 1.12 | $9.28 \times 10^{-2}$ | 1.01 | 0.91, 1.13 | $8.07 \times 10^{-1}$ | 2.77 | $2.51 \times 10^{-1}$ | 27.7 |
| ***Ethnicity*[2]** | | | | | | | | | | | | |
| Non-Hispanic White | – | | – | – | | – | – | | – | – | – | – |
| Asian or Pacific Islander | 0.8 | 0.67, 0.96 | $1.36 \times 10^{-2}$ | 1.4 | 1.08, 1.81 | $1.04 \times 10^{-2}$ | 1.45 | 0.93, 2.26 | $9.69 \times 10^{-2}$ | 15.13 | $5.18 \times 10^{-4}$ | 86.8 |
| Hispanic | 0.89 | 0.70, 1.14 | $3.63 \times 10^{-1}$ | 1.1 | 0.74, 1.62 | $6.51 \times 10^{-1}$ | 0.97 | 0.39, 2.40 | $9.40 \times 10^{-1}$ | 0.81 | $6.66 \times 10^{-1}$ | 0 |
| Non-Hispanic Black | 0.89 | 0.83, 0.96 | $1.97 \times 10^{-3}$ | 0.95 | 0.85, 1.06 | $3.23 \times 10^{-1}$ | 1.26 | 1.06, 1.50 | $9.22 \times 10^{-3}$ | 13.15 | $1.40 \times 10^{-3}$ | 84.8 |
| **Obesity[3]** | 1.18 | 1.13, 1.23 | $1.84 \times 10^{-14}$ | 0.96 | 0.90, 1.02 | $2.14 \times 10^{-1}$ | 1.03 | 0.92, 1.16 | $6.01 \times 10^{-1}$ | 29.95 | $3.14 \times 10^{-7}$ | 93.3 |
| **Adenoma Onset Age (yrs)[4]** | | | | | | | | | | | | |
| 0-49 | 1.17 | 1.11, 1.23 | $6.77 \times 10^{-9}$ | 0.67 | 0.61, 0.73 | $3.42 \times 10^{-19}$ | 0.93 | 0.81, 1.06 | $2.56 \times 10^{-1}$ | 113.24 | $< 2 \times 10^{-16}$ | 98.2 |
| ≥50 | – | - | – | – | - | – | – | - | – | – | – | – |
| **Family History of Polyps[5]** | 1.04 | 0.93, 1.17 | $4.64 \times 10^{-1}$ | 1.33 | 1.12, 1.58 | $1.12 \times 10^{-3}$ | 1.01 | 0.74, 1.38 | $9.64 \times 10^{-1}$ | 5.83 | $5.42 \times 10^{-2}$ | 65.7 |
| **Family History of Colorectal Cancer[6]** | 1.24 | 1.18, 1.31 | $4.44 \times 10^{-16}$ | 1.15 | 1.06, 1.25 | $8.09 \times 10^{-4}$ | 1.09 | 0.94, 1.26 | $2.42 \times 10^{-1}$ | 4.18 | $1.24 \times 10^{-1}$ | 52.2 |

Model adjustments: [1]adjusted for ethnicity; stratified by age groups; [2]adjusted for sex; stratified by age group; [3]adjusted for sex and ethnicity; stratified by age group; [4]adjusted for sex, ethnicity, use of Non-steroidal anti-inflammatory drugs (NSAIDs), use of Aspirin, family history of colorectal cancer, and family history of polyps; stratified by obesity and age group; [5]adjusted for sex, ethnicity, use of NSAIDs, use of Aspirin, and family history of colorectal cancer; stratified by obesity, Adenoma Onset Stratification, and age group; [6]adjusted for sex, ethnicity, use of NSAIDs, use of Aspirin, and family history of polyps; stratified by obesity, Adenoma Onset Stratification, and age group. Abbreviations: HR = Hazard Ratio, CI = Confidence Interval

**eTable 2.** Adjusted Hazard Ratios for Adenoma Recurrence by Adenoma Characteristics Across Time Periods.

| *Variables* | < 5 Years | | | 5-10 Years | | | > 10 Years | | | Heterogeneity | | |
|---|---|---|---|---|---|---|---|---|---|---|---|---|
| | HR | 95% CI | *P*-Value | HR | 95% CI | *P*-Value | HR | 95% CI | *P*-Value | Cochran's Q | *P*-value | $I^2$ (%) |
| ***Adenoma Characteristics*[¥]** | | | | | | | | | | | | |
| ***Adenoma Types*[1]** | | | | | | | | | | | | |
| Tubular | – | - | – | – | - | – | – | - | – | – | – | – |
| Villous | 2.55 | 2.31, 2.81 | $4.55 \times 10^{-79}$ | 0.9 | 0.77, 1.04 | $1.44 \times 10^{-1}$ | 2.36 | 1.89, 2.94 | $2.65 \times 10^{-14}$ | 133.22 | $1.18 \times 10^{-29}$ | 98.5 |
| Tubulovillous | 2.18 | 1.99, 2.39 | $1.58 \times 10^{-61}$ | 0.55 | 0.46, 0.66 | $1.60 \times 10^{-10}$ | 0.75 | 0.54, 1.05 | $9.07 \times 10^{-2}$ | 198.04 | $9.93 \times 10^{-44}$ | 99 |
| Serrated | 1.3 | 1.18, 1.44 | $1.10 \times 10^{-7}$ | 0.9 | 0.79, 1.03 | $1.14 \times 10^{-1}$ | 0.99 | 0.73, 1.34 | $9.52 \times 10^{-1}$ | 19.65 | $5.41 \times 10^{-5}$ | 89.8 |
| Not defined | 0.53 | 0.48, 0.57 | $1.02 \times 10^{-54}$ | 0.6 | 0.56, 0.65 | $5.16 \times 10^{-34}$ | 0.76 | 0.66, 0.88 | $1.93 \times 10^{-4}$ | 18.08 | $1.19 \times 10^{-4}$ | 88.9 |
| Mixed | 1.37 | 1.30, 1.45 | $6.99 \times 10^{-32}$ | 0.77 | 0.72, 0.82 | $2.89 \times 10^{-14}$ | 0.93 | 0.80, 1.07 | $3.01 \times 10^{-1}$ | 180.63 | $5.99 \times 10^{-40}$ | 98.9 |
| ***Dysplasia*[2]** | | | | | | | | | | | | |
| Low grade | – | - | – | – | - | – | – | - | – | – | – | – |
| High grade | 2.86 | 2.54, 3.22 | $4.13 \times 10^{-69}$ | 0.64 | 0.46, 0.90 | $9.74 \times 10^{-3}$ | 0.63 | 0.31, 1.28 | $1.99 \times 10^{-1}$ | 81.49 | $2.02 \times 10^{-18}$ | 97.5 |
| ***Number of Polyps*[3]** | | - | | | - | | | - | | | | |
| <3 | – | - | – | – | - | – | – | - | – | – | – | – |
| ≥3 | 1.69 | 1.60, 1.79 | $5.95 \times 10^{-80}$ | 1 | 0.91, 1.10 | $9.93 \times 10^{-1}$ | 0.99 | 0.80, 1.23 | $9.18 \times 10^{-1}$ | 99.72 | $2.22 \times 10^{-22}$ | 98 |
| Not defined | 1.12 | 1.04, 1.21 | $3.81 \times 10^{-3}$ | 1.58 | 1.46, 1.71 | $8.35 \times 10^{-30}$ | 1.57 | 1.38, 1.77 | $1.21 \times 10^{-12}$ | 44.21 | $2.51 \times 10^{-10}$ | 95.5 |
| ***Adenoma Size (mm)*[4]** | | | | | | | | | | | | |
| <10 | – | - | – | – | - | – | – | - | – | – | – | – |
| ≥10 | 1.63 | 1.53, 1.73 | $5.72 \times 10^{-57}$ | 0.78 | 0.70, 0.86 | $3.51 \times 10^{-6}$ | 0.9 | 0.73, 1.11 | $3.43 \times 10^{-1}$ | 158.44 | $3.93 \times 10^{-35}$ | 98.7 |
| Not Defined | 0.79 | 0.75, 0.83 | $6.94 \times 10^{-20}$ | 0.98 | 0.93, 1.04 | $5.53 \times 10^{-1}$ | 1.05 | 0.94, 1.17 | $3.97 \times 10^{-1}$ | 41.41 | $1.02 \times 10^{-9}$ | 95.2 |
| ***Adenoma Type*** | | | | | | | | | | | | |
| Low Risk | – | - | – | – | - | – | – | - | – | – | – | – |
| High Risk | 2.79 | 2.68, 2.91 | $2.22 \times 10^{-16}$ | 1.16 | 1.09, 1.24 | $1.02 \times 10^{-5}$ | 1.51 | 1.33, 1.71 | $3.07 \times 10^{-10}$ | 534.97 | $6.79 \times 10^{-117}$ | 99.6 |

[¥]Cox proportional hazards models with time-dependent effects, adjusted for sex, ethnicity, use of nonsteroidal anti-inflammatory drugs, use of aspirin (NSAIDs), family history of colorectal cancer, and family history of polyps; models 1 to 4 additionally adjusted for other polyp-related variables (polyp type, dysplasia, number of polyps, size of polyps [mm]) when not the primary variable. All models are stratified by age group; however, models 1 to 4 are additionally stratified by obesity and polyp onset stratification. Abbreviations: HR = Hazard Ratio, CI = Confidence Interval.

**eTable 3.** Adjusted Hazard Ratios for Adenoma Recurrence by Adenoma Characteristics Across Ethnicities for 5 year follow-up.

| Variables | Non-Hispanic White | | | | Asian or Pacific Islander | | | | Hispanic | | | | Non-Hispanic Black | | | | |
|---|---|---|---|---|---|---|---|---|---|---|---|---|---|---|---|---|---|
| | **HR** | **HR Lower** | **HR Upper** | ***P*-value** | **HR** | **HR Lower** | **HR Upper** | ***P*-value** | **HR** | **HR Lower** | **HR Upper** | ***P*-value** | **HR** | **HR Lower** | **HR Upper** | ***P*-value** | **Co** |
| **Adenoma Characteristics**[¥] | | | | | | | | | | | | | | | | | |
| ***Adenoma Types***[1] | | | | | | | | | | | | | | | | | |
| Tubular | - | - | - | - | - | - | - | - | - | - | - | - | - | - | - | - | |
| Villous | 2.68 | 2.41 | 2.98 | $5.02 \times 10^{-73}$ | 3.13 | 1.43 | 6.84 | $4.29 \times 10^{-3}$ | 2.69 | 0.62 | 11.66 | $1.87 \times 10^{-1}$ | 2.42 | 1.73 | 3.39 | $2.49 \times 10^{-7}$ | |
| Tubulovillous | 2 | 1.81 | 2.22 | $6.04 \times 10^{-41}$ | 1.96 | 0.87 | 4.43 | $1.06 \times 10^{-1}$ | 2.55 | 1.1 | 5.91 | $2.93 \times 10^{-2}$ | 1.77 | 1.38 | 2.28 | $6.99 \times 10^{-6}$ | |
| Serrated | 1.3 | 1.17 | 1.44 | $6.24 \times 10^{-7}$ | 0.41 | 0.13 | 1.35 | $1.43 \times 10^{-1}$ | 0.43 | 0.1 | 1.87 | $2.63 \times 10^{-1}$ | 1.37 | 0.95 | 1.97 | $8.83 \times 10^{-2}$ | |
| Not defined | 0.58 | 0.53 | 0.65 | $3.94 \times 10^{-25}$ | 0.42 | 0.24 | 0.74 | $2.33 \times 10^{-3}$ | 0.36 | 0.17 | 0.77 | $8.53 \times 10^{-3}$ | 0.51 | 0.4 | 0.63 | $3.20 \times 10^{-9}$ | |
| Mixed | 1.34 | 1.27 | 1.42 | $4.72 \times 10^{-24}$ | 1 | 0.65 | 1.52 | $9.84 \times 10^{-1}$ | 0.83 | 0.45 | 1.54 | $5.59 \times 10^{-1}$ | 1.26 | 1.07 | 1.48 | $5.94 \times 10^{-3}$ | |
| ***Dysplasia***[2] | | | | | | | | | | | | | | | | | |
| Low grade | – | – | – | – | – | – | – | – | – | – | – | – | – | – | – | – | |
| High grade | 2.25 | 1.97 | 2.56 | $9.91 \times 10^{-35}$ | 4.9 | 2.16 | 11.12 | $1.46 \times 10^{-4}$ | 3.39 | 1.27 | 9.04 | $1.46 \times 10^{-2}$ | 2.84 | 2.05 | 3.93 | $3.73 \times 10^{-10}$ | |
| ***Number of Polyps***[3] | | | | | | | | | | | | | | | | | |
| <3 | - | - | - | - | - | - | - | - | - | - | - | - | - | - | - | - | |
| ≥3 | 1.54 | 1.45 | 1.63 | $2.60 \times 10^{-46}$ | 2.27 | 1.47 | 3.5 | $2.23 \times 10^{-4}$ | 0.57 | 0.26 | 1.23 | $1.53 \times 10^{-1}$ | 1.67 | 1.42 | 1.98 | $1.85 \times 10^{-9}$ | |
| Unknown | 1.27 | 1.15 | 1.4 | $2.67 \times 10^{-6}$ | 1.61 | 0.97 | 2.66 | $6.51 \times 10^{-2}$ | 0.87 | 0.42 | 1.8 | $7.16 \times 10^{-1}$ | 1.21 | 0.97 | 1.52 | $9.16 \times 10^{-2}$ | |
| ***Adenoma Size***[4] | | | | | | | | | | | | | | | | | |
| <10 | - | - | - | - | - | - | - | - | - | - | - | - | - | - | - | - | |
| ≥10 mm | 1.51 | 1.42 | 1.61 | $3.66 \times 10^{-36}$ | 1.49 | 0.88 | 2.5 | $1.35 \times 10^{-1}$ | 1.92 | 0.94 | 3.94 | $7.32 \times 10^{-2}$ | 1.36 | 1.12 | 1.65 | $1.92 \times 10^{-3}$ | |
| Unknown | 0.87 | 0.82 | 0.93 | $1.39 \times 10^{-5}$ | 0.93 | 0.63 | 1.38 | $7.31 \times 10^{-1}$ | 0.99 | 0.56 | 1.74 | $9.61 \times 10^{-1}$ | 0.93 | 0.8 | 1.09 | $3.77 \times 10^{-1}$ | |
| **Adenoma Type** | | | | | | | | | | | | | | | | | |
| High Risk | 1.42 | 1.29 | 1.56 | $8.37 \times 10^{-13}$ | 1.55 | 1.06 | 2.26 | $2.32 \times 10^{-2}$ | 1.01 | 0.61 | 1.69 | $9.62 \times 10^{-1}$ | 1.41 | 1.2 | 1.65 | $3.31 \times 10^{-5}$ | |
| Low Risk | - | - | - | - | - | - | - | - | - | - | - | - | - | - | - | - | |

[¥]Cox proportional hazards models with time-dependent effects, adjusted for sex, ethnicity, use of nonsteroidal anti-inflammatory drugs, use of aspirin (NSAIDs), family history of colorectal cancer, and additionally adjusted for other polyp-related variables (polyp type, dysplasia, number of polyps, size of polyps [mm]) when not the primary variable. All models are stratified by age group; however, mo obesity and polyp onset stratification. Abbreviations: HR = Hazard Ratio, CI = Confidence Interval.

**eTable 4:** Adjusted Hazard Ratios for Adenoma Recurrence by Adenoma Characteristics Across Sex for 5 year follow-up.

| Variables | Female | | | | Male | | | | Heterogeneity | | |
|---|---|---|---|---|---|---|---|---|---|---|---|
| | **HR** | **HR Lower** | **HR Upper** | ***P*-value** | **HR** | **HR Lower** | **HR Upper** | ***P*-value** | **Cochran's Q** | ***P*-value** | **$I^2$ (%)** |
| **Adenoma Characteristics**[¥] | | | | | | | | | | | |
| ***Adenoma Types***[1] | | | | | | | | | | | |
| Tubular | – | – | – | – | – | – | – | – | – | – | – |
| Villous | 2.74 | 2.4 | 3.12 | $3.81 \times 10^{-51}$ | 2.59 | 2.24 | 3 | $1.37 \times 10^{-37}$ | 0.32 | $5.74 \times 10^{-1}$ | 0 |
| Tubulovillous | 2.21 | 1.93 | 2.53 | $2.67 \times 10^{-30}$ | 1.8 | 1.59 | 2.04 | $1.29 \times 10^{-20}$ | 4.78 | $2.88 \times 10^{-2}$ | 79.1 |
| Serrated | 1.36 | 1.2 | 1.55 | $2.41 \times 10^{-6}$ | 1.19 | 1.02 | 1.39 | $2.44 \times 10^{-2}$ | 1.7 | $1.92 \times 10^{-1}$ | 41.1 |
| Not defined | 0.59 | 0.53 | 0.66 | $1.08 \times 10^{-18}$ | 0.55 | 0.49 | 0.62 | $7.17 \times 10^{-25}$ | 0.73 | $3.92 \times 10^{-1}$ | 0 |
| Mixed | 1.43 | 1.32 | 1.55 | $3.19 \times 10^{-19}$ | 1.24 | 1.15 | 1.33 | $3.75 \times 10^{-9}$ | 6.65 | $9.90 \times 10^{-3}$ | 85 |
| ***Dysplasia***[2] | | | | | | | | | | | |
| Low grade | - | - | - | - | - | - | - | - | – | – | – |
| High grade | 2.43 | 2.03 | 2.91 | $2.33 \times 10^{-22}$ | 2.3 | 1.97 | 2.68 | $4.77 \times 10^{-26}$ | 0.21 | $6.49 \times 10^{-1}$ | 0 |
| ***Number of Polyps***[3] | | | | | | | | | | | |
| <3 | - | - | - | - | - | - | - | - | – | – | – |
| ≥3 | 1.64 | 1.51 | 1.78 | $2.04 \times 10^{-33}$ | 1.48 | 1.38 | 1.59 | $8.63 \times 10^{-26}$ | 3.44 | $6.38 \times 10^{-2}$ | 70.9 |
| Unknown | 1.27 | 1.14 | 1.42 | $1.70 \times 10^{-5}$ | 1.27 | 1.14 | 1.42 | $1.87 \times 10^{-5}$ | 0 | $1.00 \times 10^{-0}$ | 0 |
| ***Adenoma Size (mm)***[4] | | | | | | | | | | | |
| <10 | - | - | - | - | - | - | - | - | – | – | – |
| ≥10 | 1.6 | 1.46 | 1.74 | $2.77 \times 10^{-25}$ | 1.42 | 1.31 | 1.54 | $1.73 \times 10^{-17}$ | 3.84 | $4.99 \times 10^{-2}$ | 74 |
| Unknown | 0.89 | 0.83 | 0.96 | $1.74 \times 10^{-3}$ | 0.87 | 0.81 | 0.94 | $2.42 \times 10^{-4}$ | 0.18 | $6.69 \times 10^{-1}$ | 0 |
| **Adenoma Type** | | | | | | | | | | | |
| High Risk | 1.49 | 1.34 | 1.65 | $4.54 \times 10^{-14}$ | 1.34 | 1.21 | 1.49 | $1.78 \times 10^{-8}$ | 2 | $1.58 \times 10^{-1}$ | 49.9 |
| Low Risk | - | - | - | - | - | - | - | - | - | - | |

[¥]Cox proportional hazards models with time-dependent effects, adjusted for sex, ethnicity, use of nonsteroidal anti-inflammatory drugs, use of aspirin (NSAIDs), family history of colorectal cancer, and family history of polyps; models 1 to 4 additionally adjusted for other polyp-related variables (polyp type, dysplasia, number of polyps, size of polyps [mm]) when not the primary variable. All models are stratified by age group; however, models 1 to 4 are additionally stratified by obesity and polyp onset stratification. Abbreviations: HR = Hazard Ratio, CI = Confidence Interval.

**Supplement 1. Pathology Report Extraction**

This protocol outlines the methodology for extracting polyp characteristics from pathology reports to characterize polyp type, number, size, and dysplasia for analysis in the Polypectomy Cohort study. Pathology reports were identified using specific keywords to ensure relevance, while unrelated notes were excluded to minimize noise. Extracted variables were structured in JSON format for consistency and downstream analysis.

**Keywords Used for Identification:**

- polypectom
- adenoma
- tubul
- villous
- sessile
- plastic
- serrated

**Exclusion Criteria (to Filter Unrelated Notes):**

- Discharge
- Re:
- Anesthesia
- Appointment
- PROBLEM LIST
- Endoscopy Nurse Note
- Consent - Surgery/Procedure
- Patient Instructions

**Extracted Variables:**

1. **Adenoma Type & Histological Type:** Categorized as:
    - Adenomatous: Tubular, Villous, Tubulovillous
    - Serrated Polyps: Hyperplastic, Sessile serrated, Traditional serrated, Unclassified serrated adenomas
    - Non-neoplastic: Inflammatory, Hamartomatous
    - Mixed Type: Multiple polyp types

2. **Number of Polyps:** Multiple fragments in one position or specimen counted as 1 polyp.
3. **Maximum Size of Largest Adenoma or Fragment:** Measured in millimeters (mm).
4. **Dysplasia Type:** Classified as Low-grade, High-grade, or "Not specified" if unspecified.

Output the result in Json format without explanation

Two Examples:

```
{
    "histological_type" : "Adenomatous: Villous",
    "number_of_polyp" : 1,
    "max_size_of_polyp" : "12 mm",
    "dysplasia_type" : "High-grade"
}

{
    "histological_type" : "Mixed Type: Tubular, Sessile serrated",
    "number_of_polyp" : 3,
    "max_size_of_polyp" : "10 mm",
    "dysplasia_type" : "Low-grade"
}
```

"
